# Artificial intelligence and opioid use: a narrative review

**DOI:** 10.1101/2022.05.18.22275269

**Authors:** Seema Gadhia, Georgia C. Richards, Tracey Marriott, James Rose

## Abstract

**Background:** Opioids are strong pain medications that can be essential for acute pain. However, opioids are also commonly used for chronic conditions and illicitly where there are well recognised concerns about the balance of their benefits and harms. Technologies using artificial intelligence (AI) are being developed to examine and optimise the use of opioids. Yet, this research has not been synthesised to determine the types of AI models being developed and the application of these models.

**Methods:** We aimed to synthesise studies exploring the use of AI in people taking opioids. We searched three databases: the Cochrane Database of Systematic Reviews, EMBASE, and Medline on 4 January 2021. Studies were included if they were published after 2010, conducted in a real-life community setting involving humans, and used AI to understand opioid use. Data on the types and applications of AI models were extracted and descriptively analysed.

**Results:** Eighty-one articles were included in our review, representing over 5.3 million participants and 14.6 million social media posts. Most (93%) studies were conducted in the USA. The types of AI technologies included natural language processing (46%) and a range of machine learning algorithms, the most common being random forest algorithms (36%). AI was predominately applied for the surveillance and monitoring of opioids (46%), followed by risk prediction (42%), pain management (10%), and patient support (2%). Few of the AI models were ready for adoption, with most (62%) being in preliminary stages.

**Conclusions:** Many AI models are being developed and applied to understand opioid use. However, there is a need for these AI technologies to be externally validated and robustly evaluated to determine whether they can improve the use and safety of opioids.

**SUMMARY BOX:** **Key Points**
Across the landscape of opioid research, natural language processing models (46%) and ensemble algorithms, particularly random forest algorithms (36%), were the most common types of AI technologies studied.
There were four domains to which AI was applied to assess the use of opioids, including surveillance and monitoring (46%), risk prediction (42%), pain management (10%), and patient support (2%). The AI technologies were at various stages of development, validation, and deployment, with most (62%) models in preliminary stages, 11% required external validation, and few models were openly available to access (6%).

## INTRODUCTION

Opioids are pain medicines related to opium that are deemed essential by the World Health Organisation (WHO).[1] There are over 200 different types of opioids that can be prescribed, purchased over-the-counter (e.g. at pharmacies), purchased online or obtained illicitly.[2–4] There are also various conditions that opioids can be used for, including but not limited to, cancer pain, postoperative pain, chronic non-cancer pain, opioid dependence and withdrawal.[5–9] Despite being essential, opioids can cause a number of adverse events from minor (e.g. constipation, nausea) to severe (e.g. addiction, depression, sleep problems),[10] as well as death.

In the USA, 128 lives were lost every day to opioid overdoses in 2018.[11] Such opioid-related deaths, widely described as the US opioid epidemic, were linked to the increased prescribing of opioids, opioid misuse, and the transition to illicit substances. In the UK, six deaths per day due to a drug overdose involved an opioid in 2018.[12] The prescribing of opioids in primary care more than doubled in England between 1998 and 2016.[13] Thus, interventions are being developed to examine and improve the use of opioids.

Several approaches have been trialled to improve the use of opioids with partial success. These have included: educational resources;[14] non-pharmacological therapies, for example, cognitive behavioural therapy, hypnosis, relaxation techniques, mindfulness, acupuncture, and exercise[15]); the monitoring of prescribing data,[16] and toolkits to support the review and safe reduction of opioids.[17] However, technological advances could help streamline such approaches to improve the use of opioids.

The use of artificial intelligence (AI) technologies in preventative health and medicines optimisation is gaining traction. For example, AI is being used to predict sudden death in heart failure patients and support the selection of appropriate treatments.[18] A review on the use of AI interventions to aid in opioid use disorders found 29 unique interventions.[19] However, this review only examined the grey literature, which is not quality checked by publishers and peer review.[20] Grey literature can include documents such as reports and online content that can provide insight into emerging research. However, the types of AI reported in peer-reviewed literature and across other aspects of opioid research has not been synthesised.

The UK published a National AI strategy in September 2021 [21] and is encouraging the use of AI to drive digital transformation across the National Health Service (NHS) [22]. NHS organisations (e.g. NHSX) are supporting the acceleration of AI technologies through financial awards[23] and the UK’s overprescribing review has recommended the commission of digital tools to tackle and reduce overprescribing.[24] However, there are few reviews that examine the use of AI to inform such policies. Therefore, the aim of this review was to explore the use of AI technologies across the landscape of opioid research.

## METHODS

We designed a narrative review to understand how AI technologies have been used, applied, and implemented in research on opioid use.

### Search strategy

An information specialist designed and ran the search strategy in three databases: the Cochrane Database of Systematic Reviews, EMBASE, and Medline. Search terms relating to “opioids” and “artificial intelligence” were included (see Table S1 in Appendix 1 for the complete list of terms). The search was initially performed on 27 January 2020 and updated on 4 January 2021. We also searched Google Scholar on 18 November 2020.

### Eligibility criteria

Studies were included if they were published after 2010, had been conducted in a real-life setting in human beings, and tested a form of AI to optimise or understand opioid use. AI was defined as ‘computer systems that can perform tasks normally requiring human intelligence’,[25] which could include any form of machine learning, deep learning, neural networks, and natural language processing. Studies were not restricted by outcomes or settings, and conference abstracts were included to capture all emerging research. Studies were excluded if they were not published in English, did not specifically relate to both opioids and AI, were conducted outside of a real-life setting, for example, in research settings exploring genes, receptor subtypes and modulators, and were not original research. Editorials and commentary were excluded.

### Study selection

Titles and abstracts were screened independently using the prespecified eligibility criteria by one review author (SG), followed by the full-text articles. Where the conference abstract and full article were both available, the full article was included.

### Data extraction

One review author (SG) extracted data from included studies into a predeveloped spreadsheet. This included: year of study and author names; country; study design; study population and data source; sample size; technology investigated; area of application; outcomes; and stage of development.

### Data analysis

The findings were descriptively synthesised by identifying areas of commonality in terms of the AI technology used and the area of application. The types of AI technology were classified based on the method described by Brownlee in the Tour of Machine Learning Algorithms.[26] The stages of development were defined based on reported findings in the studies and categorised into seven groups: preliminary research; model development required; model development planned; external validation required; prototype for scale-up developed; local implementation; openly available.

## RESULTS

We screened 389 titles and abstracts and 118 full texts for eligibility (Figure 1). There were 81 studies that met our eligibility criteria and were included in the review (Table 1). Of the 81 studies, 18 were conference abstracts, which are summarised in Table S2 Appendix 2.

**Figure 1.**
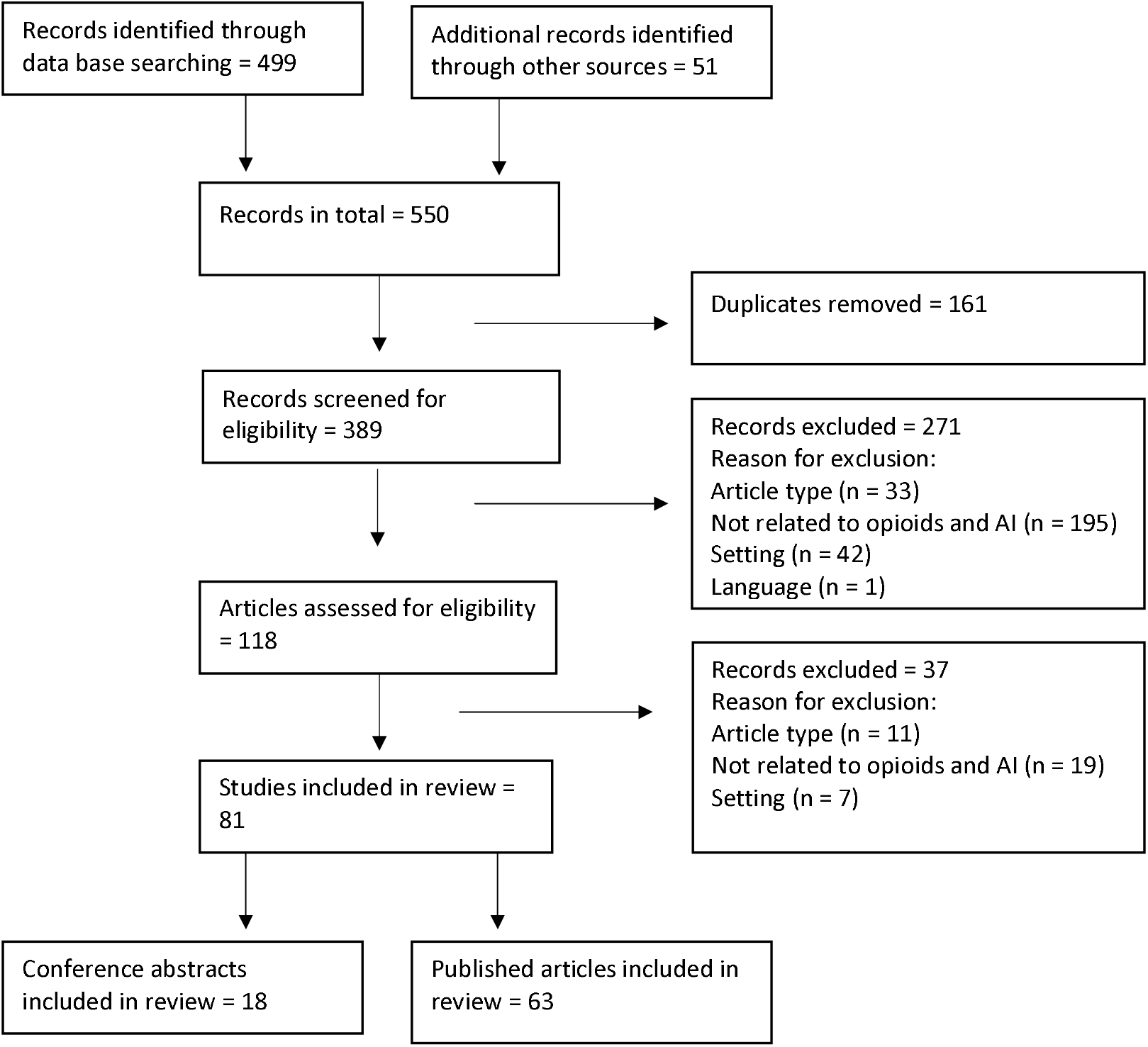
Flow diagram of the study selection

**Table 1.** Summary of the 63 included studies on artificial intelligence and opioid use that were published in academic journals, ordered alphabetically by the four domains to which AI was applied, including surveillance and monitoring, risk prediction, pain management, and patient support.

| Study ID (country)[ref] | Study design | Study population and data source | Sample (n=) | AI Technology | Application | Outcomes | Stage of development |
| --- | --- | --- | --- | --- | --- | --- | --- |
| **Risk prediction** |  |  |  |  |  |  |  |
| Ahn et al. 2016 (Bulgaria). [27] | Cross-sectional | Amphetamine or heroin-dependent or polysubstance dependent adults. Data collected from two four-hour study sessions using a battery of self-reported & administered assessments. | 222 | Machine learning (elastic net) | To identify substance-specific behavioural markers for heroin and amphetamine dependence. | The machine-learning approach revealed substance-specific multivariate profiles that classified heroin and amphetamine dependence. Psychopathy was uniquely associated with heroin dependence. | Preliminary research |
| Anderson et al. 2020 (USA). [28] | Prognostic | Military patients (males) undergoing a specific orthopaedic procedure. Data from the Military Health System Data Repository. | 10,919 | Logistic regression, random forest, Bayesian belief network, and gradient boosting machine models | Risk prediction of prolonged opioid use after a specific orthopaedic procedure. | The gradient boosting machine can be used to understand factors contributing to opiate misuse after ACL reconstruction. The most influential features with a positive association for prolonged opioid use are preoperative morphine equivalents, pharmacy ordering site locations, shorter deployment time, and younger age. | Local implementation and undergoing external validation. |
| Ben-Ari et al. 2017 (USA). [29] | Retrospective cohort | Male patients in the Veterans Affairs system who had TKA. Data from EHRs. | 32,636 | Natural language processing-based machine-learning classifier | Assessment of the association of long-term opioid use on adverse outcomes after TKA. | The accuracy of the text classifier was 0.94 with an AUROC of 0.99. Long-term opioid use prior to TKA was associated with an increased risk of knee revision during the first year after TKA among predominantly male patients. | Preliminary research. |
| Boslett et al. 2020 (USA). [30] | Retrospective cohort | People with an unclassified drug overdose recorded in death records. Data from the National Centre for Health Statistics Detailed Multiple Cause of Death records. | 632,331 | Random forest ensemble | Comparison of methodologies to predict the involvement of opioids in unclassified drug overdose deaths to estimate the number of fatal opioid overdoses. | Random forest models performed similarly to logistic regression. Using a superior prediction model, the study found that 71.8% of unclassified drug overdoses in 1999–2016 involved opioids, approximately 28% more than reported. There was geographic variation in undercounting of opioid overdoses. | Preliminary research. |
| Calcaterra et al. 2018 (USA). [31] | Retrospective cohort | All patients discharged from Denver Health Medical Centre, an integrated safety net health system. Data from EHRs. | 27,705 | Random forest, least absolute shrinkage, and selection operator (lasso), stepwise logistic regression | Prediction of COT in hospitalised patients not on opioids before hospitalisation. | The multiple logistic regression model correctly predicted 79% of the COT patients and 78% of the no COT patients. Accuracy was 78%, and the AUROC was 0.86. Risk factors for COT included more than 10 mg of morphine equivalents prescribed per day during hospitalisation, two or more opioid prescriptions filled in the year preceding hospitalisation, past year receipt of non-analgesic pain medications, and past year receipt of benzodiazepines. | External validation required. |
| Che et al. 2017 (USA). [32] | Retrospective cohort | Patients who received at least one opioid prescription Data from EHRs. | 102,166 | Deep feed-forward neural network, recurrent neural network, logistic regression, support vector machine Random forest | Classification of opioid users. | The deep learning models were able to achieve superior classification performance and identify useful feature indicators for opioid-dependent and long-term users. Several disorders diagnoses, such as “substance-related disorders”, “anxiety disorders”, and “other mental health disorders” are all highly related to opioid dependence. | Model development planned. |
| Dong et al. 2020 (USA). [33] | Retrospective cohort | Patients with at least one historic encounter before first opioid poisoned related diagnosis. Data from EHRs. | 555,000 | Deep neural networks Random forest, decision tree Logistic regression | Risk prediction of future opioid poisoning and the most important features for such predictions. | EHR based prediction can achieve best recall with random forest method (85.7%), best precision with deep learning (precision). Top predictive feature with regards diagnosis is ‘sedative, hypnotic or anxiolytic dependence, continuous’. The top predictive factors in the health facts dataset are pulse and heart rate. | Model development planned. |
| Ellis et al. 2019 (USA). [34] | Case-control | Patients diagnosed with substance misuse. Data from EHRs. | 7,797 | Random forest | Prediction of substance dependence based on lab tests and vital signs | The top machine learning classifier using all features achieved a mean AUROC of ~ 92%. The study found opioid dependent patients have significantly higher WBC and respiratory disturbances. Opioid dependent patients are also commonly malnourished which is characterized by low red cell distribution width, and blood albumin compared to controls. | Preliminary research |
| Green et al. 2019 (USA). [35] | Retrospective cohort | Patients who had overdosed on an opioid medication. Data from EHRs. | 977 | NLP | Identification and classification of opioid-related overdoses. | Code-based algorithms developed to detect opioid-related overdoses and classify them according to heroin involvement performed well. The NLP-enhanced algorithms for suicides/ suicide attempts and abuse-related overdoses perform significantly better than code-based algorithms and are appropriate for use in settings that have data and capacity to use NLP. | Model development required. |
| Haller et al. 2017 (USA). [36] | Retrospective cohort | Patients with chronic non-cancer pain. Data from EHRs. | 3,668 | NLP | Prediction of opioid abuse by use of automated risk assessments. | Confirmed through manual review, the NLP algorithm had 96.1% sensitivity, 92.8% specificity, and 92.6% positive predictive value in identifying opioid agreement violation. Patients classified as high risk were three times more likely to violate opioid agreements compared with those with low/moderate risk. | External validation required. |
| Han et al. 2020 (USA). [37] | Cross-sectional | Adolescents. Self-reported data collected from a survey. | 41,579 | Neural networks, distributed, random forest, gradient boosting machine model | Prediction of opioid misuse | The overall rate of opioid misuse among adolescents was 3.7% (n=1521). Prediction performance was similar across the four models AUROC values range from 0.809 to 0.815). In terms of the area under the precision-recall curve (AUPRC), the distributed random forest showed the best performance in prediction (0.172). | Preliminary research. |
| Hastings et al. 2020 (USA). [38] | Retrospective cohort | Patients prescribed opioid medication. Data from EHRs. | 80,768 | Regularized regression, neural network | Prediction of future opioid dependence, abuse or poisoning in advance of an initial opioid prescription. | All models achieve an AUC near 0.800, indicating they have strong predictive power but could still be improved. The two variables with the largest odds ratios (indicating increased risk) are related to crime: | Preliminary research. |
|  |  |  |  |  |  | release from prison and an indicator for an arrest. Individuals released from prison in the prior year are estimated as 119% more likely to develop an adverse outcome if given an initial prescription, all else equal, and those with an arrest in the prior year are 76% more likely to do so. |  |
| Hylan et al. 2015 (USA). [39] | Retrospective cohort | Patients with chronic non-cancer pain initiating opioid therapy. Data from EHRs. | 2,752 | NLP | Prediction of risk for problem opioid use in a primary care setting. | The AUROC (c-statistic) for problem opioid use was 0.739. As predictive models for problem opioid use are only moderately accurate, at best, there is always a need for the clinician's vigilance to ensure safe and appropriate opioid use for long-term management of chronic musculoskeletal pain. | Preliminary research. |
| Karhade et al. 2019A (USA). [40] | Case-control | Patients undergoing anterior cervical discectomy and fusion for degenerative disorders. Data from EHRs. | 2,737 | Random forest, stochastic gradient boosting, neural network, support vector machine, elastic net penalized, logistic regression | Prediction of sustained opioid prescription after anterior cervical discectomy and fusion | The stochastic gradient boosting algorithm achieved the best performance (c-statistic 0.81). Global explanations of the model demonstrated that preoperative opioid duration, antidepressant use, tobacco use, and Medicaid insurance were the most important predictors of sustained postoperative opioid prescription. | External validation required. |
| Karhade et al. 2019B (USA). [41] | Retrospective cohort | Patients undergoing total hip arthroplasty for osteoarthritis. Data from EHRs. | 5,507 | Stochastic gradient boosting, random forest, support vector machine, neural network, elastic net penalized logistic regression | Prediction of sustained postoperative opioid prescriptions after total hip arthroplasty | The elastic net penalized logistic regression model achieved the best performance (c-statistic 0.77). 6.3% of patients had prolonged postoperative opioid prescriptions. The factors determined for prediction of prolonged postoperative opioid prescriptions were age, duration of opioid exposure, preoperative haemoglobin, and preoperative medications (antidepressants, benzodiazepines, nonsteroidal anti-inflammatory drugs, and beta-2-agonists). | External validation required. |
| Karhade et al. 2019C (USA). [42] | Case-control | Patients undergoing surgery for lumbar disc herniation. Data from EHRs. | 5,413 | Random forest, stochastic gradient boosting, neural network, support vector machine, elastic-net penalized logistic regression | Prediction of prolonged opioid prescription after surgery for lumbar disc herniation | 7.7% of patients were identified, with sustained postoperative opioid prescription after surgery. The elastic-net penalized logistic regression model had the best overall performance (c-statistic 0.81). The three most important predictors were: instrumentation, duration of preoperative opioid prescription, and comorbidity of depression. | Available online as open access. |
| Karhade et al. 2020A (USA). [43] | Retrospective cohort | Opioid naïve adults who underwent lumbar spine surgery. Data from EHRs. | 8,435 | Random forest, stochastic gradient boosting, neural network, support vector machine, elastic-net penalized logistic regression | Prediction of prolonged opioid prescriptions in opioid-naïve lumbar spine patients | 4.3% of patients were found to have prolonged postoperative opioid prescriptions. The elastic-net penalized logistic regression achieved the best performance (c-statistic=0.70). The five most important factors for prolonged opioid prescriptions were use of instrumented spinal fusion, preoperative benzodiazepine use, preoperative antidepressant use, preoperative gabapentin use, and uninsured status. | Available online as open access. |
| Katakam et al. 2020B | Retrospective cohort | Patients undergoing surgery for total knee replacement. | 12,542 | Random forest, stochastic gradient | Preoperative prediction of prolonged opioid | The stochastic gradient boosting model had the best performance. Age, history of preoperative opioid use, | External validation required. |
| (USA). [44] |  | Data from EHRs. |  | boosting, neural network, support vector machine, elastic-net penalized logistic regression | prescriptions after total knee replacement. | marital status, diagnosis of diabetes, and several preoperative medications were predictive of prolonged postoperative opioid prescriptions. |  |
| Lo-Ciganic et al. 2019 (USA). [45] | Prognostic | Patients without cancer receiving one or more opioid prescription. Data from prescription drug and medical claims. | 560,057 | Multivariate logistic regression, least absolute shrinkage and selection operator-type regression, random forest, gradient boosting machine, deep neural network | Prediction of opioid overdose risk | The deep neural network (c statistic = 0.91) and gradient boosting machine (c statistic = 0.90) algorithms outperformed the other methods for predicting opioid overdose. The deep neural network classified patients into low-risk (76.2% of the cohort), medium-risk (18.6% of the cohort), and high-risk (5.2% of the cohort) subgroups, with only 1 in 10 000 in the low-risk subgroup having an overdose episode. More than 90% of overdose episodes occurred in the high-risk and medium-risk subgroups. | Preliminary research. |
| Lo-Ciganic et al. 2020A (USA). [46] | Prognostic | Patients without cancer receiving one or more opioid prescription. Data from prescription drug and medical claims. | 361,527 | Elastic net, random forests, gradient boosting machine, deep neural network | Prediction of incident of OUD diagnosis. | All approaches had similar prediction performances (c-statistic ranged from 0.874 to 0.882); elastic net required the fewest predictors. This algorithm was able to segment the population into different risk groups based on predicted risk scores, with 70% of the sample having minimal OUD risk, and half of the individuals with OUD captured in the top decile group. | Preliminary research. |
| McCann-Pineo et al. 2020 (USA). [47] | Retrospective cohort | Patients ≥18 years who had an ED visit. Data from survey. | 44,227 | LASSO regularization, elastic net regularization, conditional inference random forest, gradient boosted machine, Naïve Bayes | Prediction of opioid administration during an ED visit and prescribing upon discharge. | The strongest predictors of ED opioid prescription were CT scan ordered, abdominal pain and back pain. Tooth pain and fracture injury diagnoses were the strongest predictors of a discharge opioid prescription. | Preliminary research. |
| Segal et al. 2020 (USA). [48] | Retrospective cohort | Patients who had made medical insurance claims. Data from commercial claims database. | 550,000 | NLP, gradient boosting machine | Prediction of early diagnosis of OUD. | The c-statistic for the model was 0.959. Significant differences between positive OUD- and negative OUD-controls were in the mean annual amount of opioid use days, number of overlaps in opioid prescriptions per year, mean annual opioid prescriptions, and annual benzodiazepine and muscle relaxant prescriptions. The new algorithm offers a mean 14.4-month reduction in time to diagnosis of OUD. | Preliminary research. |
| Wadekar et al. 2020 (USA). [49] | Retrospective cohort | Adults responding to the National Survey on Drug Use and Health (2016). | 42,324 | Random forest | Prediction for risk for opioid use disorder and identify interactions between various characteristics that increase this risk. | Random forest predicted adults at risk for OUD with the average AUROC over 0.89. Initiation of marijuana before 18 years emerged as the dominant predictor. Early marijuana initiation increased the risk if individuals were between 18–34 years, or had incomes less than \$49,000, or were of Hispanic and White heritage, or were on probation, or lived in neighbourhoods with easy access to drugs. | Preliminary research. |

| Surveillance and monitoring |  |  |  |  |  |  |  |
| --- | --- | --- | --- | --- | --- | --- | --- |
| Afshar et al. 2019 (USA). [50] | Prognostic | Patients with opioid misuse who had an inpatient hospitalisation. Data from EHRs. | 6,224 | NLP, topic modelling (Latent Dirichlet Allocation) | Identification of subtypes of patients with opioid misuse and examining the distinctions between the subtypes. | Distinct subtypes were identified after examining data and applying methods in artificial intelligence. Class 1 patients had high hospital utilisation with known opioid-related conditions (36.5%); Class 2 included patients with illicit use, low socioeconomic status, and psychoses (12.8%); Class 3 contained patients with alcohol use disorders with complications (39.2%); and class 4 consisted of those with low hospital utilisation and incidental opioid misuse (11.5%). | External validation required. |
| Anwar et al. 2020 (USA). [51] | Retrospective observational | Opioid-related Twitter posts relating to prescription opioids, heroin, and synthetic. | 10,000 posts | NLP | Investigation of the extent to which the content of opioid-related tweets corresponds with the triphasic nature (shift from prescription opioids for pain, to heroin and then to synthetic opioids) of the opioid crisis and correlates with opioid overdose deaths. | Tweets were classified as relating to prescription opioids, heroin, and synthetic opioids using NLP. The pattern of opioid-related Twitter posts resembled the triphasic nature of the opioid crisis. Tweets mentioning heroin and synthetic opioids were significantly associated with opioid overdose deaths. | Preliminary research. |
| Badger et al. 2019 (USA). [52] | Retrospective cohort | Patients with an ICD-9 and ICD-10 code related to opioid overdose and poisoning. Data from EHRs. | 278 | NLP, Naïve Bayes, support vector machine, LASSO logistic regression, random forest | Development of machine learning models for classifying the severity of opioid overdose events. | Random forest models using features derived from a common data model and free text can be effective for classifying opioid overdose events. Key word features extracted using NLP such as 'Narcan' and 'Endotracheal Tube' are important for classifying overdose event severity. | External validation required. |
| Black et al. 2020 (USA). [53] | Retrospective cohort | People who died of a drug poisoning. Data from surveillance system program. | 4,008 | NLP | Assessment of changes in mortality rates in ER/LA opioid analgesics after the implementation of the Risk Evaluation and Mitigation Strategy (REMS). | The NLP model correctly identified all active pharmaceutical ingredients with 100% sensitivity and specificity relative to what was printed on the death certificate. The population-adjusted mortality rate of ER/LA opioid analgesics has decreased after the implementation of the REMS in three states. | Preliminary research. |
| Cai et al. 2020 (USA). [54] | Retrospective infoveillance | Indiana geolocated tweets filtered for geocoded messages in the immediate pre and post period of the HIV outbreak. | 5,112 posts | Unsupervised machine learning approach using NLP called the Biterm Topic Model | Identification and characterization of tweets related to the 2015 Indiana HIV outbreak. | The Biterm Topic Model identified 1350 tweets thought to be relevant to the outbreak and then confirmed 358 tweets using human annotation. The most prevalent themes identified were tweets related to self-reported abuse of illicit and prescription drugs, OUD, self-reported HIV status, and public sentiment regarding the outbreak. Geospatial analysis found that these messages clustered in population dense areas outside of the outbreak. | Preliminary research. |
| Carrell et al. 2015 (USA). [55] | Retrospective cohort | Patients receiving chronic opioid therapy, a mixed-model health plan. Data from EHRs. | 22,142 | NLP | Identification of problem opioid use from clinical notes of patients receiving chronic | Traditional diagnostic codes for problem opioid use were found for 2240 (10.1%) patients. NLP-assisted manual review identified an additional 728 (3.1%) | Model development required. |
|  |  |  |  |  | opioid therapy. | patients with evidence of clinically diagnosed problem opioid use in clinical notes. |  |
| Chary et al. 2017 (USA). [56] | Retrospective observational | Tweets that contained at least one keyword related to prescription opioid use. | 3,611,528 posts | NLP | Correlation of geographic variation of social media posts mentioning prescription opioid misuse with government estimates of misuse. | Natural language processing can be used to analyse social media to provide insights for syndromic toxico-surveillance. Mention of misuse of prescription opioids (MUPO) on Twitter correlate strongly with state-by-state National Surveys on Drug Usage and Health (NSDUH) estimates of MUPO. The strongest correlation occurred between data from Twitter and NSDUH data from those aged 18–25. | Model development required. |
| Cuomo et al. 2020 (USA). [57] | Retrospective observational | Tweets related to opioid, heroin/injection, and HIV behaviour. | 1,350 posts | Unsupervised machine learning approach using NLP (Biterm Topic Model) | Identification and characterisation of HIV outbreak triggered by opioid abuse and transition to injection drug use. | Prevalent themes identified were tweets related to self-reported abuse of illicit and prescription drugs, opioid use disorder, self-reported HIV status, and public sentiment regarding the outbreak. Geospatial analysis found messages clustered in population dense areas outside of the outbreak. | Preliminary research. |
| Fodeh et al. 2021 (USA). [58] | Retrospective observational | Tweets containing key opioid related keywords. | 1,677 | NLP, recurrent neural networks, random forest, support vector machines | Categorisation of twitter chatter based on the motive of opioid misuse. | A recurrent neural network classifier (XLNet) achieved the best performance. The model identified three groups of tweets: tweets of users with no OM, tweets of users with pain-related OM and tweets of users with recreational-related OM. Clinically, individuals who misuse opioids because of pain have different motivations and patterns of use. | Preliminary research. |
| Hazelhurst et al. 2019 (USA). [59] | Retrospective cohort | Patients who had overdosed on opioid medication. Data from EHRs. | 305 | NLP | Identification and classification of opioid-related overdoses. | The method performed well in identifying overdose, intentional overdose, and involvement of opioids (excluding heroin) and heroin. The method performed poorly at identifying adverse drug reactions and overdose due to patient error and fairly at identifying substance abuse in opioid-related unintentional overdose. | Preliminary research. |
| Jha & Singh 2019 (USA). [60] | Retrospective observational | Recreational drug use reddit, and drug addiction recovery reddit. | 170,097 | NLP and machine learning (SMARTS software) | Identification of individuals open to addiction recovery interventions using SMARTS. SMARTS is a public, open source, web-based application for addiction information extraction, analysis, and modelling. | SMARTS generalises well across the different kinds of addiction posts and can identify individuals open to recovery interventions for intoxicants, such as opioids with an accuracy of 96%. The SMARTS web server and source code are available at: http://haddock9.sfsu.edu/. | Available online as open access. |
| Jungquist et al. 2019 (USA). [61] | Retrospective observational | Patients undergoing back, neck, hip, or knee surgery. Data from enrolled patients at a community hospital. | 60 | Support vector machine | Development of machine learning to aid in earlier detection of respiratory depression. | The model provides a high detection rate (>0.9) when the detection horizon is short. However, even for longer time horizons (e.g., 10 min before the actual event), the model that uses all three electronic measurements is able to correctly predict nearly 80% of opioid-induced respiratory depression events. Nurses can use electronic monitoring data to identify patients experiencing OIRD to influence opioid-sparing pain management. | Preliminary research. |
| Kalyanam et al. 2017 (USA). [62] | Retrospective observational | Tweets filtered for commonly abused prescription opioid drugs. | 11 million posts | Biterm topic model | Identification of macro non-medical use of prescription opioid medication | The cluster purity for each drug was up to three times better than that of a random set of tweets. Twitter content was associated with a high degree of discussion (approximately 80%) about polydrug abuse involving multiple types of substances. | Preliminary research. |
| Khemani et al. 2017 (USA). [63] | Retrospective cohort | Patients presenting with abdominal pain to the emergency department. Data from EHRs. | 16,121 | NLP | Characterisation of opioid use, constipation, and risk factors for surgical diagnoses among non-cancer patients presenting with acute abdominal pain (AAP). | Approximately 19% of adults presenting with AAP were opioid users; constipation is almost 3 times as likely in opioid users compared to non-opioid users presenting with AAP. Age and neutrophil count independently predicted increased risk, and chronic opioid use decreased risk of surgical diagnosis. | Preliminary research. |
| Li et al. 2019 (USA). [64] | Retrospective observational | Instagram posts based on opioid keywords. | 12,857 | Recurrent neural network, random forest, decision tree, support vector machine | Identification of illegal internet drug dealing. | 1228 drug dealer posts comprising 267 unique users were detected. The deep learning model reaching 95% on F1 score and performing better than the other three models. By removing the hashtags in the text, the model had better performance. | Model development required. |
| Lingeman et al. 2017 (USA). [65] | Retrospective cohort | Primary care outpatients taking a prescribed opioid. Data from EHRs. | 112 | NLP<br>Support vector machine | Surveillance of drug-related aberrant behaviour. | The model could differentiate clinical encounter notes that contain opioid-related aberrant behaviour from those that do not with relatively high accuracy (81%). Mentions of illicit drug use and patient anxiety were strong predictors of documented aberrant behaviour. | Model development planned. |
| Mackey et al. 2017 (USA). [66] | Retrospective observational | Tweets filtered for prescription opioid keywords. | 619,937 posts | Biterm topic model | Identification of the marketing of illegal online sales of controlled substances. | The biterm topic model enabled identification of 1778 (0.003%) containing content associated with illicit online drug sales. These tweets represent a potential patient safety hazard and substance abuse risk. | Preliminary research. |
| Mackey et al. 2018 (USA). [67] | Retrospective observational | Tweets using common opioid keywords. | 213,041 | Biterm topic model | Detection and reporting illicit online pharmacy selling of controlled substances. | Using the biterm topic model, 0.32% (692/213,041) tweets were identified as being associated with illegal online marketing and sale of prescription opioids. After removing duplicates and dead links, we identified 34 unique “live” tweets, with 44% directing consumers to illicit online pharmacies, 32% linked to individual drug sellers, and 21% used by marketing affiliates. In addition to offering the “no prescription” sale of opioids, many of these vendors also sold other controlled substances and illicit drugs. | Prototype for potential scale-up developed. |
| Masters et al. 2018 (USA). [68] | Case-control | Patients who received COT. Data from EHRs. | 11,253 | NLP | Associated health care costs of COT patients with POU. | COT patients with NLP-identified, manually validated POU experienced significantly higher costs than COT patients without POU in the first year following their index date. The greatest difference in costs was observed around the time of identification of POU. The difference was driven by large differences in resource utilisation in the 30 days following clinician labelling of | Preliminary research. |
|  |  |  |  |  |  | POU. |  |
| Mojtabai et al. 2019 (USA). [69] | Retrospective cohort | Adult participants using opioids in the past year. Data was self-reported from a national survey on drug use. | 30,813 | Boosted regression | Assessment of the prevalence and correlates of self-reported misuse of prescribed opioids. | In boosted regression analysis, misuse of prescription opioids without a prescription, misuse of prescribed benzodiazepines, other substance use disorders, illegal activities, and psychological distress were the most influential factors associated with prescribed opioid misuse. | Preliminary research. |
| Palmer et al. 2015 (USA). [70] | Retrospective cohort | Patients receiving COT. Data from EHRs. | 22,142 | NLP | Prevalence of POU. | Agreement between the NLP methods and ICD-9 coding was moderate (kappa 0.61). 9.4% of COT patients had current problem opioid use, with higher rates observed among young COT patients, patients who sustained opioid use for more than 4 quarters, and patients who received higher opioid doses. | Preliminary research. |
| Panlilio et al. 2020 (USA). [71] | Retrospective cohort | Methadone-maintained participants undergoing contingency-management treatment. Retrospective data from three randomised clinical trials. | 309 | Unsupervised machine learning | Patterns of opioid and cocaine use in contingency management, methadone-treated participants. | Four clusters of use patterns were identified, which can be described as opioid use, cocaine use, dual use (opioid and cocaine), and partial/complete abstinence. Contingency management increased membership in clusters with lower levels of drug use and fewer symptoms of substance use disorder. | Preliminary research. |
| Paulose et al. 2018 (India). [72] | Retrospective observational | Tweets containing the keyword fentanyl. | 4,604 | NLP | Identification of fentanyl misuse using social media posts. | The sentiment analysis algorithm labelled 610 (13.25 %) tweets as positive. Crisis, dead, death, die, dose, drug, heroin, kill, lethal, opioid, overdose and police were some of the words frequently associated with fentanyl. There was a high correlation and association of fentanyl with these negative terms which demonstrated fentanyl abuse in the real world. | Preliminary research. |
| Prieto et al. 2020 (USA). [73] | Retrospective cohort | Patients treated for opioid misuse (OM) by paramedics. Data from Denver Health paramedic trip reports. | 54,359 | Random forest, k-nearest neighbours, support vector machines, L1-regularized logistic regression | Identification of potential OM from paramedic documentation. | L1-regularized logistic regression was the highest performing algorithm (AUROC=0.94) in identifying OM. Among trip reports with reviewer agreement, 77.79% (907/1166) were considered to include information consistent with OM. | Preliminary research. |
| Sarker et al. 2019A (USA). [74] | Retrospective observational | Tweets that mentioned prescription and illicit opioids. | 9,006 | Support vector machines, random forest, deep convolutional neural network | Monitoring of population-level opioid abuse. | Deep convolutional neural networks marginally outperformed support vector machines and random forests, with an accuracy of 70.4%. Geolocation data was able to identify the origins of tweets at the state level, it may be possible to further narrow down to the county or city level in the future. | Model development planned. |
| Sarker et al. 2019B (USA). [75] | Retrospective observational | Tweets that mentioned prescription and illicit opioids. | 9,006 | NLP, Naive bayes decision tree, k-nearest neighbours, random forest, support vector machine, deep convolutional neural network | Development of an automatic text-processing pipeline for geospatial and temporal analysis of opioid-mentioning social media chat. | Ensemble of 4 classifiers (Ensemble_1: CNN, RF, SVM, NB) producing the best F1 score (0.726). 19.4% of tweets were related to abuse, 22.2% were related to information, 53.6% were unrelated, and 4.7% were not in English. Yearly rates of abuse-indicating social media post showed statistically significant correlation with county-level opioid-related overdose death rates for 3 years. | Model development planned. |
| Sharma et al. 2020 (USA). [76] | Retrospective cohort | Adult inpatients at a hospital and tertiary academic centre. Data from EHRs. | 1,000 | Logistic regression, convolutional neural network | Development of a PHI free model for text classification of opioid misuse. | The top performing models with AUROCs > 0.90 included concept unique identifier codes as inputs to a convolutional neural network, max pooling network, and logistic regression model. The model demonstrates good test characteristics for an opioid misuse computable phenotype that is void of any PHI and performs similarly to models that use PHI. | Available via Github. |
| Singh et al. 2019 (USA). [77] | Case series | Patients who were brought to the ED while undergoing naloxone treatment following an opioid overdose. Data from a wearable biosensor. | 11 | Random forest, adaboost, support vector machine, logistic regression | Development of a wearable biosensor to detect collaborative non-adherence detection during opioid abuse surveillance. | The best performing algorithm was the Random Forest with an AUROC = 0.93. Overall, we achieved an average detection accuracy of 90.96% when the collaborator was one of the patients in the dataset, and 86.78% when the collaborator was from a set of users unknown to the classifier. | Model development planned. |
| Sinha et al. 2017 (USA). [78] | Retrospective cohort | Patients treated with chronic pain medication. Data from EHRs. | 212,343 | NLP | Demographic patterns of opioid dependent patients in New York. | The trends of opioid dependence among the clinic population indicate that the prevalence is more in a certain section of the population. The predominance is among the non-Hispanic, white population in 19 to 38 year olds. | Preliminary research. |
| Yao et al. 2019 (USA). [79] | Case-control | Posts of suicidality among opioid users on Reddit. | 45,459 posts | Convolutional neural network, logistic regression, random forest, support vector machines | Detection of suicidality among opioid users on Reddit | Best classifier was convolutional neural network, which obtained an F1 score of 96.6% and AUC of 0.932. When predicting out-of-sample data for posts containing both suicidal ideation and signs of opioid addiction, neural network classifiers produced more false positives and traditional methods produced more false negatives. | Preliminary research. |
| **Pain management** |  |  |  |  |  |  |  |
| Goyal et al. 2020 (USA). [80] | Cross-sectional | Children (<18 years) who presented to the ED with a long bone fracture. Data from EHRs of 7 paediatric EDs. | 8,533 | NLP | To identify if minority children with long-bone fractures are less likely to receive analgesics; receive opioid analgesics, and achieve pain reduction | NLP identified patients with radiology reports indicating long bone fractures. Minority children are more likely to receive analgesics and achieve 2-point reduction in pain, however, they are less likely to receive opioids and achieve optimal pain reduction. | Preliminary research. |
| Gram et al. 2017 (Germany) [81] | Retrospective cohort | Patients admitted to hospital for a total hip replacement. Data were clinical parameters recorded from patients the day prior to surgery. | 81 | Support vector machine | Preoperative identification of risk factors for analgesic inefficacy of postoperative opioid treatment. | The accuracy of the model was 65 %. Severity of the pre-surgical chronic pain condition was a factor associated with post-surgical insufficiency of analgesic treatment. It was possible to predict analgesic efficacy based on the pre-operative EEG recordings using machine learning, with similar accuracy to the chronic pain grade. | Preliminary research. |
| Graves et al. 2018 (USA). [82] | Retrospective observational | Patients and caregivers' reviews that contained brand, generic, or colloquial name of an opioid. Data from all Yelp reviews of hospital. | 836 | NLP | Characterisation of patients and caregivers' reviews about pain management and opioids. | Themes identified in natural language processing of opioid reviews with five-star and one-star ratings reflected pain management and opioid-related themes similar to those identified by manual coding. Yelp reviews describing experiences with pain management and opioids had lower ratings compared with other reviews. Negative descriptions of pain management and opioid-related experiences were more commonly described than positive experiences, and the number of themes they reflected was more diverse. | Preliminary research. |
| Gudin et al. 2020 (USA). [83] | Cohort | Chronic pain patients. Data was self-reported in questionnaires. | 127 | Hybrid combining multi-objective optimization and support vector regression | Reducing opioid prescriptions by identifying responders on topical analgesic treatment. | The model can predict the outcomes with accuracy of AUROC between 73.8–87.2% and this allowed their incorporation in a decision support system for the selection of the treatment of chronic pain patients. | External validation required. |
| Lee et al. 2021 (USA). [84] | Retrospective cohort | Patients undergoing total joint replacement surgery. Data from EHRs and a patient survey conducted at a non-profit community hospital. | 285 | Random forest, XGBoost, logistic regression, support vector machine, k-nearest neighbours (K-NN), neural network models | Identification of patients who may need less or more opioids after being discharged from TJR surgeries. | XGBoost and Random Forest models achieve the best test accuracy of 83% (AUROC 0.72;0.65). A machine learning classification model was developed that can identify patients expected to use less opioids and to detect opioid overusers within two weeks after undergoing total joint replacement surgeries. | Preliminary research. |
| Nair et al. 2020 (USA). [85] | Retrospective cohort | Adults ($\geq 18$ years) undergoing ambulatory surgery. Data from institution's information management system data warehouse. | 13,700 | Multinomial regression, naïve Bayesian, neural network, random forest, extreme gradient boosting | Prediction of post-operative opioid requirements for pain management of ambulatory surgery patients. | The best performing model, the random forest, showed that the lower opioid requirements are predicted with better accuracy (89%) as compared with higher opioid requirements (43%). The type of procedure, medical history and procedure duration were the top features contributing to model predictions. Overall, the contribution of patient and procedure features towards model predictions were 65% and 35% respectively. | External validation required. |
| Pantano et al. 2020 (Italy). [86] | Retrospective cohort | Adults diagnosed with cancer with stable background pain in the last week. Data from the Italian Oncologic Pain Survey. | 4,016 | Un-supervised machine learning | Identification of clinical features for breakthrough cancer pain (BTcP) and differential opioid response. | The algorithm identified 12 distinct BTcP clusters. Optimal BTcP opioids-to-basal pain opioids ratios differed across the clusters, ranging from 15% to 50%. The optimal dose of BTcP opioids depended on the dose of basal opioids. | Available online as open access. |
| Parthipan et al. 2019 (USA). [87] | Retrospective cohort | Patients receiving surgery at a tertiary care academic medical centre with symptoms of depression. Data from EHRs. | 430 | NLP, elastic net regularized regression | Postoperative pain management in depressed patients | The NLP algorithm identified depression with an F1 score of 0.95. Patients receiving an SSRI and opioid prodrug had significantly worse pain control at discharge, 3 and 8-week follow up. Preoperative pain, surgery type, and opioid tolerance were the strongest predictors of pain control. The study results imply direct acting opioids (e.g., oxycodone or morphine) may be better choices for depressed patients on SSRIs for pain management. | Preliminary research. |
| **Patient support** |  |  |  |  |  |  |  |
| Epstein et al. 2020 (USA). [88] | Retrospective cohort | Participants seeking treatment for OUD at a treatment | 189 | Random forest | Individual patient level prediction of stress and drug | Models achieved overall accuracy of 0.93 by the end of 16 weeks of tailoring. This was driven mostly by correct | Model development planned. |
|  |  | research clinic. Data was from self-reported questionnaires, physical examinations, and psychological testing. |  |  | craving ninety minutes in the future with passively collected GPS data. | predictions of absence. For predictions of presence, “believability” (positive predictive value, PPV) usually peaked in the high 0.70s toward the end of the 16 weeks. When target events are comparatively subtle, like stress or drug craving, accurate detection or prediction probably needs effortful input from users, not passive monitoring alone. |  |
| Scherzer et al. 2020 (USA). [89] | Pilot study using a prospective cohort design & qualitative interview | Patients receiving buprenorphine treatment for OUD at an outpatient clinic. Data collected through the mobile App and interviews. | 40 | NLP | Development of mobile peer support for patients with OUD. | The Marigold App will allow patients to access a tailored support group 24/7 and is augmented with AI tools capable of understanding the emotional sentiment in messages, automatically “flagging” critical or clinically relevant content. | Model development planned. |

AI: artificial intelligence; AUROC: area under the receiver operating characteristic curve; COT: chronic opioid therapy; EHR: electronic health records; ER: extended-release; LASSO: Least Absolute Shrinkage and Selection Operator; LA: long-acting; NLP: Natural language processing; OM: opioid misuse; OUD: opioid use disorder; POU: problem opioid use; PHI: Protected Health Information; TKA: total knee arthroplasty

The included studies represented over 5.3 million participants and 14.6 million social media posts. The majority (93%, n=75) of studies were conducted in the USA with the remainder being performed in Bangladesh (n=1), Bulgaria (n=1), Germany (n=1), India (n=1), Israel (n=1) and Italy (n=1). Of the published articles (n=63), most studies used observational designs, including cohort (54%), retrospective observational (22%), case-control (8%), prognostic (6%), and cross-sectional (5%). One study was a case series, one was a pilot study using mixed methods, and there was one retrospective infovelliance study (Table 1). The main sources of data for testing AI models were medical records and claims databases (54%) and social media (20%) (Figure 2).

**Figure 2.**
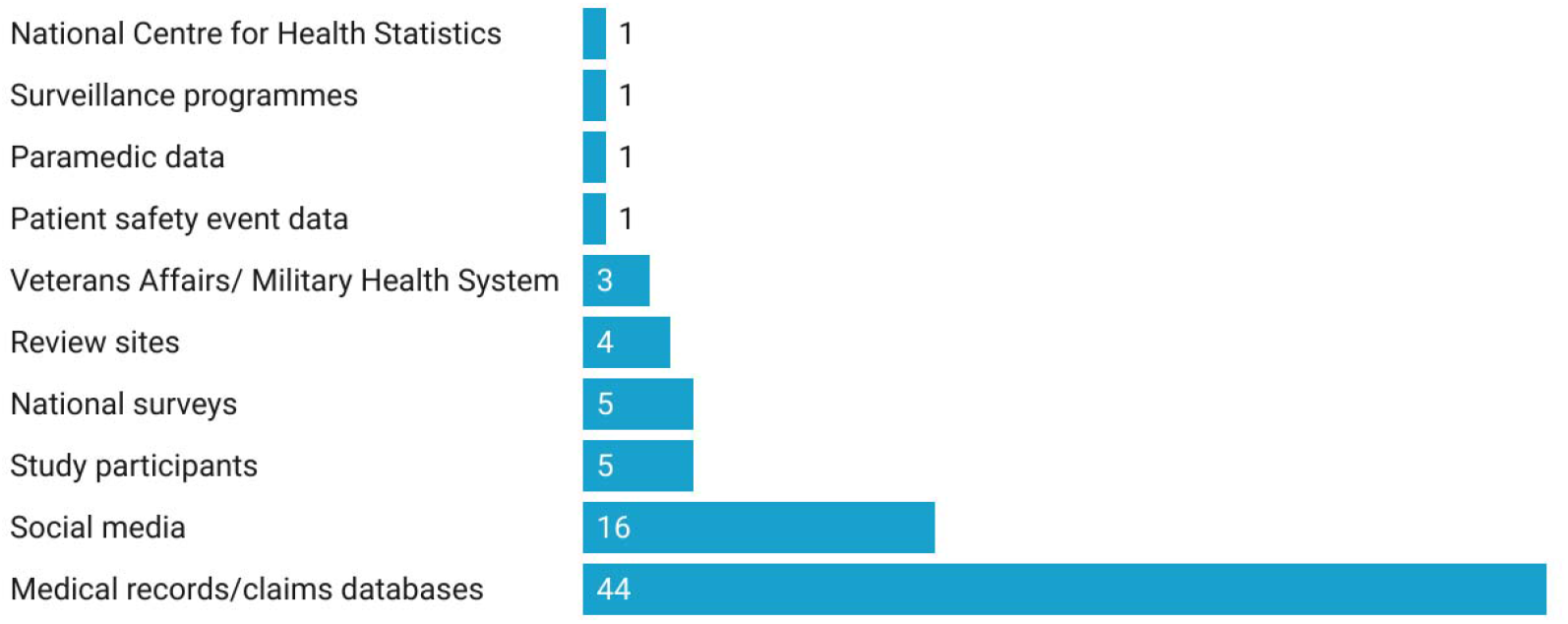
Data Sources of all included studies (n=81) for AI opioid research

The areas where AI technology was being applied and tested broadly fell into four distinct categories: surveillance and monitoring of activity or consequences such as misuse, respiratory depression, and HIV outbreaks (46%, n=37); risk prediction of outcomes such as opioid use disorder, dependence, or overdose (42%, n= 34); pain management (10%, n=8); and patient support technology (2%, n=2) (Appendix 3). For studies that focused on risk prediction (n= 34), 18% specifically investigated prolonged opioid use following surgery (Table 2).

**Table 2:** Predictors of risk for a defined adverse outcome following opioid use in AI research, reported in 34 included studies

| Adverse outcome | Number of studies (%) | Variable predictive of risk |
| --- | --- | --- |
| Prolonged opioid use after surgery | 7 (21) | Age; marital status; preoperative opioid use and duration; preoperative medications (antidepressants, benzodiazepines, nonsteroidal anti-inflammatory drugs, gabapentin, and beta-2-agonists); medications commonly used to treat anxiety and insomnia; preoperative haemoglobin; tobacco use; comorbidity of depression or diabetes; instrumentation; Medicaid insurance and particular pharmacy ordering sites. |
| Opioid use disorder | 4 (12) | Mean annual amount of opioid use days; number of overlaps in opioid prescriptions per year; mean annual opioid prescriptions; annual benzodiazepine and |
|  |  | muscle relaxant prescriptions; initiation of marijuana before 18 years; pain; mental health issues; traumatic brain injury; and male gender. Dynamics through time-in-treatment of decision-making parameters and symptom intensity (craving, anxiety, and withdrawal symptoms). |
| Opioid dependence | 3 (9) | Psychopathy; higher WBC and respiratory disturbances; malnutrition, and reduced sensitivity to loss. |
| Opioid poisoning and overdose | 3 (9) | Sedative, hypnotic, or anxiolytic dependence; arrest history; the number of overdoses in a person's social network; early refills; total days' supply; concomitant use of antidepressants; concomitant use of antipsychotics; total opioid claims and high-dose opioid-benzodiazepine use. |
| Opioid abuse and misuse | 2 (6) | Violation of opioid agreements; release from prison and an indicator for an arrest. |
| Chronic opioid therapy | 1 (3) | More than 10 mg of morphine equivalent/per day during hospitalisation; two or more opioid prescriptions filled in the year preceding the index hospitalisation; past year receipt of non-analgesic pain medications and past year receipt of benzodiazepines. |
| Emergency Department opioid prescription | 1 (3) | CT scan ordered, abdominal pain and back pain. |

Ensemble algorithms (59%, n=48), particularly random forest algorithms (36%, n=29) and natural language processing models (46%, n=37) were the most common types of AI technology researched (Table 3). In terms of efficacy measures, several studies (43%, n=35) used the area under the receiver-operating characteristic (AUROC) curve. Other efficacy measures used included the macro averaged F1 score, Brier score, positive predictive value, and negative predictive value.

**Table 3:** AI technologies used in opioid research by all included studies (n=81), based on Brownlee’s Tour of Machine Learning Algorithms.[26]

| **AI technology** | **Number of studies\*** |
| --- | --- |
| ***Ensemble algorithms*** | ***48*** |
| Random forest | 29 |
| Gradient boosting machine | 7 |
| Stochastic gradient boosting | 5 |
| eXtreme Gradient Boosting | 4 |
| Boosted tree | 1 |
| Adaboost | 1 |
| Ensemble (unspecified) | 1 |
| ***Natural Language Processing*** | ***37*** |
| Natural language processing | 29 |
| Natural language processing and the Biterm Topic Model | 5 |
| Natural language processing and machine learning | 2 |
| Natural language processing and topic modelling (Latent Dirichlet Allocation) | 1 |
| ***Deep Learning Algorithms*** | ***23*** |
| Neural network | 10 |
| Deep neural network | 5 |
| Convolution neural network | 4 |
| Recurrent neural network | 4 |
| ***Instance-based algorithms*** | ***22*** |
| Support vector machine | 18 |
| K-Nearest Neighbour | 4 |
| ***Regularization algorithms*** | ***16*** |
| Elastic Net | 10 |
| Least absolute shrinkage and selection operator (LASSO) | 6 |
| ***Regression algorithms*** | ***13*** |
| Logistic regression | 10 |
| Regularized regression | 1 |
| Multinomial regression | 1 |
| Boosted regression | 1 |
| **Bayesian algorithms** | **5** |
| Naïve Bayes | 4 |
| Bayesian belief network | 1 |
| **Decision tree algorithms** | **4** |
| Decision tree | 4 |
| **Clustering algorithms** | **1** |
| bi-kmeans clustering modelling | 1 |
| **Other** | **1** |
| Hybrid combining multi-objective optimization and support vector regression | 1 |

*\*some studies reported the use of multiple types of AI technologies in a single study thus the total number is greater than 81*

The AI models included in the review were at various stages of development, validation, and deployment. The majority (62%, n=50) were at the preliminary stage, 11% (n=9) required external validation, few models were openly available to access (6%, n=5) (Figure 3).

**Figure 3.**
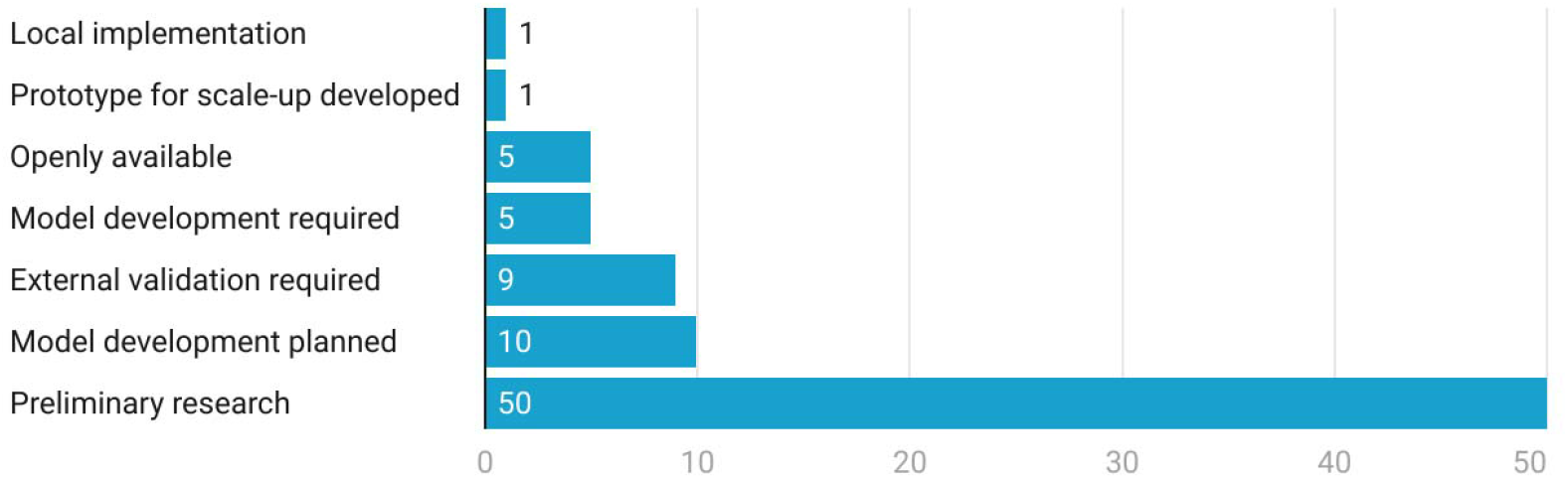
Stage of development of the AI models in opioid research from all included studies (n=81)

## DISCUSSION

We identified 81 studies that tested AI in people using opioids. Most research was from the USA, with no studies reported in the UK. While the opioids themselves presented similar risks, the populations studied were very different. There were various types of AI models being used, including a range of machine learning algorithms and models using natural language processing. The majority of included studies examined the use of AI in risk prediction and surveillance and monitoring, two areas that could have significant patient safety benefits to prevent long-term opioid use and opioid dependence.

Most of the studies reviewed focused on developing AI technology to support the identification of factors that could predict the increased risk of developing an adverse opioid-related outcome. The need for pre-operative identification of people at risk of sustained opioid use following surgery has been recommended by experts.[90] Thus, this is a potential gap that AI technology could fill if studies were robustly designed.

A commonly reported predictive factor across nearly all studies on risk prediction was the concomitant use of sedative and anxiolytic medication such as benzodiazepines. An evidence review by Public Health England identified an increased risk of long-term opioid use in people who had previously used or were currently using benzodiazepines.[91] A systematic review into factors associated with high-dose opioids reported that people co-prescribed benzodiazepines is a high priority area for targeted interventions and coordinated strategies.[92]

Progress of AI development in detecting illegal use also has the potential to significantly reduce the number of opioid-related deaths related to misuse. However, to enable effective interventions to be developed in this area also requires localised real-time intelligence. AI technology focused on detecting illegal use combined with real-time intelligence could support agencies that are currently working in this area to target activity at the time it is detected.

Guidelines and educational resources have been developed that support clinicians with pain management and highlight the problems associated with long term opioid treatment.[14,93] To date, these have had limited impact on improving opioid prescribing, with prescribing of high doses of opioids in some areas showing an increasing trend.[16] However, AI technology to improve pain management used in combination with educational resources and guidelines could be a way to prevent unnecessary escalation and long-term use of opioid treatment.

Beaulieu and colleagues have reviewed the grey literature on opioid use disorder and AI interventions, identifying 29 unique interventions.[94] Our narrative review expanded on this research to assess the AI technology across all areas of opioid research and includes a wider scope of literature. Similar to our review, Beaulieu and colleagues found a lack of scientific evaluation,[94] which was also highlighted by Hossain and colleagues in their conference abstract on AI in opioid research and practice.[95] The literature on AI models for diagnosing ischemic stroke has also illustrated the variation in measuring efficacy and the need for standardisation. [96,97]

### Strengths and limitations

Conference abstracts were included in our review to capture emerging research. However, we did not follow up on further development and validation of models beyond the reported findings in the publication. Post-publication, several of the models may have undergone further development and external validation and may now be available publicly. Thus, we were limited by the reporting of information in included studies and the infancy of AI research. In particular, it was difficult to evaluate the performance of AI models as various efficacy measures were used.

It was challenging to find a comprehensive way to classify the various types of AI technologies being researched. Various systems have been described that include classification by complexity,[98] learning style and algorithm similarity.[26] The method described by Brownlee that classified AI technology according to similarity was chosen for the review as it clearly described which algorithms fell into each category.[26] Using this system, it was found that natural language processing models and the random forest algorithm were the most commonly researched AI technology used by the studies.

Our review only included studies published in English, which could exclude published research conducted in non-English speaking countries. Finally, we conducted a narrative review that did not involve a quality assessment of the included studies. However, nearly all the studies included in this review were observational. Thus, high-quality research is required to test the efficacy and effectiveness of AI technologies in people using or receiving opioids.

### Implications

Public Health initiatives have focused on identifying and addressing people that are taking opioids long-term or becoming dependent on opioids,[91,99] yet this process has not been systematically embedded in clinical practice. Preventative interventions involving AI early in the care pathway could improve the systematic nature of such initiatives and reduce the number of people taking long-term opioids or dependant. AI technology could also support intelligence to reduce illegal and illicitly available opioids, to identify the prevalence and local hot spots to target interventions. However, before the adoption of AI models in clinical practice, future research should be conducted to standardise methods and determine whether AI models are superior to current initiatives in clinical practice.

To conduct AI research, large datasets are required. Hence, we found that electronic health records and social media posts were most often used. The NHS holds relevant records and data, on tens of millions of patients, from a huge and ethnically diverse population.[100] Limitations around curation, management and secure access to this data could be barrier to adoption of these AI advances in the UK unless developments are made.[101] Access to good quality data is also recognised as a current barrier and, vital enabler, in the UK National AI strategy.[21] The strategy makes recommendations to review datasets and their availability to support the development of AI models. However, this access to data needs to be expediated to enable AI advances to have any practical use in the UK soon.

The increased use of AI technology is a key recommendation in the NHS Long-Term Plan,[22] thus funding should be allocated to conduct randomised control trials and prospective studies in UK healthcare settings. To enable validation and implementation of AI technology into care pathways, collaboration between many stakeholders is required, including developers, healthcare organisations, clinicians, and patients. National guidance on the development, testing, and implementation of AI technologies would standardise such processes and help organisations to ensure that patients are protected.

## Conclusions

Various AI technologies have been applied to several areas of opioid use, yet this research is still in its infancy. The effectiveness of AI technologies in reducing opioid use and harms cannot be determined until robust randomised and prospective studies are conducted. Therefore, there is a clear need for these AI models to be validated and robustly evaluated. To facilitate the spread and adoption of innovation in this area, collaboration of organisations, developers, funders, researchers, prescribers, and patients is required.

## Data Availability

All data produced in the present work are contained in the manuscript and appendices

## ACKNOWLEDGEMENTS

We acknowledge Anne Gray, Knowledge Specialist at Oxford Academic Health Science Network, for carrying out the literature search.

## APPENDIX 1.

**Table S1:** Search terms

| Source | Date of search | Search terms |
| --- | --- | --- |
| MEDLINE<br>In-process<br>and other<br>non-<br>indexed<br>citations<br>and<br>MEDLINE | 04/01/21 | (((((opioid*).ti,ab OR (opiate*).ti,ab OR (narcotic*).ti,ab)<br>**AND** (("artificial intelligence").ti,ab OR (AI).ti,ab OR<br>("machine learning").ti,ab OR ("natural language<br>processing").ti,ab OR (nlp).ti,ab OR ("deep learning").ti,ab))<br>**OR**<br>(exp *"ANALGESICS, OPIOID"/ **AND** (*"ARTIFICIAL<br>INTELLIGENCE"/ OR *"COMPUTER HEURISTICS"/ OR<br>*"EXPERT SYSTEMS"/ OR *"FUZZY LOGIC"/ OR<br>*"KNOWLEDGE BASES"/ OR *"MACHINE LEARNING"/ OR<br>*"NATURAL LANGUAGE PROCESSING"/ OR *"NEURAL<br>NETWORKS, COMPUTER"/)))<br>NOT (editorial).pt) [DT 2010-2021] [Languages English]" |
| EMBASE | 04/01/21 | "((((opioid*).ti,ab OR (opiate*).ti,ab OR (narcotic*).ti,ab)<br>**AND** (("artificial intelligence").ti,ab OR (AI).ti,ab OR<br>("machine learning").ti,ab OR ("natural language<br>processing").ti,ab OR (nlp).ti,ab OR ("deep learning").ti,ab))<br>**OR**<br>(exp *"NARCOTIC ANALGESIC AGENT"/ **AND** exp<br>"ARTIFICIAL INTELLIGENCE"/))<br>NOT (editorial).pt) [DT 2010-2021] [English language]" |
| Cochrane<br>Library –<br>CENTRAL | 25/11/20 | (Exp Artificial intelligence and exp narcotics) or<br>("artificial intelligence" or AI or "natural language<br>processing" or NLP or "deep learning" or "machine<br>learning"):ti,ab,kw AND (opioid* or opiate* or<br>narcotic*):ti,ab,kw<br>Limited to 2010-2020. |

## APPENDIX 2.

**Table S2:** Summary of the 18 conference abstracts that were included in the review, ordered alphabetically by the domain to which AI was applied to assess the use of opioids, including surveillance and monitoring and risk prediction

| Study ID (country)[ref] | Data source | Sample (n=) | AI Technology | Application | Outcome | Stage of development |
| --- | --- | --- | --- | --- | --- | --- |
| **Risk prediction** |  |  |  |  |  |  |
| Crosier 2017 (USA).[102] | Opioid users. Data from enrolled opioid users. (n = 260) | 260 | Random forest | Prediction of overdose frequency and identification of key predictive features | The model performed a binary classification to predict lifetime overdose status, with an error rate of 30.25%. Arrest history and the number of overdoses in a person's social network emerged as the most important predictors of overdose. | Preliminary research |
| Li 2018 (USA).[103] | Patients prescribed opioid medication. Data from the IMS LifeLink PharMetrics PlusTM database. | 1,246,642 | Boosted tree | Prediction of opioid overdoses among prescription opioid users | The boosted tree classifier outperformed other learning algorithms (c-statistic 0.77). The most significant prognostic features were, early refills, total days' supply, concomitant use of antidepressants, concomitant use of antipsychotics, and total opioid claims. | Preliminary research |
| Lo-Ciganic 2020B (USA).[104] | Medicaid beneficiaries who had made a medical claim. Integration of human services data, criminal justice records, and medical examiner's autopsy data with medical claims data. | 79,086 | Gradient boosting machine | Prediction of risk of opioid overdose. | The gradient boosting machine algorithm including comprehensive integrated data outperformed the model using medical claims only (c-statistic=0.920). Over 85% of individuals with overdoses were in the top two deciles having the highest overdose rates. Few individuals had overdose episodes in the bottom eight deciles. | Preliminary research |
| Lopez-Guzman 2019 (USA).[105] | Patients with OUD being treated in and outpatient setting. | 74 | LASSO logistic regression | Prediction of clinical outcomes for opioid use disorder using decision-making trajectories. | Most variables did not survive LASSA regression for relapse suggesting most of these personality factors, while useful for diagnosis, are not determinants of prognosis. The dynamics through time-in-treatment of decision-making parameters and symptom intensity (craving, anxiety, and withdrawal symptoms) were significant predictors of relapse. | Preliminary research |
| None 2019 (USA).[106] | Opioid naïve and non-naïve adults undergoing surgery. Data from claims data from the Clinformatics DataMart (OptumInsight). | 199,423 | Non-linear machine learning | Prediction of postoperative opioid prescription refills. | Compared with linear models, nonlinear models led to better performance AUROC = 0.754 vs 0.738. Medications commonly used to treat anxiety and insomnia were most predictive, especially among opioid-naïve patients. These findings suggest that patient attributes, such as sleep disorders and anxiety, can be used to predict postoperative refill. | Preliminary research |
| Sim on 2018 (USA).[107] | Patients with two years of continuous insurance eligibility selected from | 23,371 | Extreme gradient boosting | Prediction of whether a patient who does not currently have OUD will receive a diagnosis in the next 12 | The model was able to predict future OUD outcomes with a surprising level of fidelity in individuals who did not have a prescription for opioids | Preliminary research |
|  | commercial and government healthcare claims databases. |  |  | months. | documented in administrative claims during the predictive period (AUROC = 0.837). |  |
| Vassileva 2019A (USA).[108] | Mono-dependent opiate and stimulant users. Data from a larger study. | 595 | Elastic net | Prediction of addiction phenotypes in mono-dependent opiate and stimulant users. | For prediction of opiate dependence, the AUROC = 0.88. Reduced sensitivity to loss in opiate users was one of the most consistent findings across different tasks and cognitive models and could represent a potential biomarker for opiate addiction. | Preliminary research. |
| Vassileva 2019B (USA).[109] | Monosubstance-dependent substance users. Data from a larger study. | 595 | LASSO penalized regression models | Identification of behavioural markers that accurately classify alcohol dependence, nicotine dependence, cannabis dependence, opiate dependence, and stimulant dependence. | For classification accuracy for opiate dependence the AUROC = 0.91. Personality variables had higher predictive utility than neurocognitive variables. Psychopathy was the only common predictor of all drug classes | Preliminary research. |
| Wang 2019 (USA). [110] | Patients newly initiated on prescription opioids. Data from inpatient or emergency department claims of fee-for service Medicaid beneficiaries. | 346 | Machine learning, specifically, bi-kmeans clustering modelling | Identification of opioid-benzodiazepine (OPI-BZD) dose and duration trajectories and subsequent opioid overdose | Opioid overdose odds varied substantially across OPI-BZD use trajectories, with individuals having consistent high-dose OPI-BZD use having more than 10 times overdose odds. | Preliminary research. |
| Weiner 2019 (USA).[111] | Adult patients who had cough and received care in a medical institution. Data from EHR in a midwestern academic medical institution. | 25,593 | NLP | Identification and characterisation of opioid-containing cough suppressants among patients with chronic cough. | About one in five patients with chronic cough received an opioid-containing cough suppressant prescription, which was more likely in this cohort than in patients with non-chronic cough. | Preliminary research. |
| Workman et al. 2019(USA).[112 ] | Patients having outpatient visits for which one or more of eight opioid prescriptions were issued. Data from EHRs. | 45,326 | Deep learning | Prediction of opioid use disorder using prescription, patient, provider, and facility characteristics predictors. | The model was able to predict OUD with an AUROC= 0.87. Pain, mental health issues, traumatic brain injury, and male gender were identified as top features in the models. New potential risk factors include respiratory, as well as behavioural health and Social Service Providers. | Model development planned. |
| **Surveillance and monitoring** |  |  |  |  |  |  |
| Carrell 2017 (USA).[113] | Patients receiving chronic opioid therapy through a staff model health care system. Data from EHR. | 15,498 | NLP and machine learning (a model based on NLP-extracted data from chart notes). | Identification of POU in large patient populations. | NLP-assisted manual review indicated that 1,453 (9.4%) patients had POU. In the validation set the algorithm achieved 56% sensitivity and 76% precision. EHR data useful for identifying with modest precision many patients experiencing POU. | Model development required. |
| Mahmud et al. 2018(USA).[114 ] | Patients admitted to hospital for a painful condition being treated with opioid analgesics. | 30 | Decision tree, K-Nearest Neighbor eXtreme Gradient Boosting | Automatic detection of opioid intake using electrodermal activity, skin temperature and triaxis acceleration data generated from a wrist worn biosensor. | Decision tree and eXtreme Gradient Boosting shows the best results. The detection the rate of the decision tree and extreme Gradient boosting is 99.3% and 99.8% respectively. | Model development planned. |
| Mazer-Amirshahi 2017 | Patients exposed to a patient safety event | 282 | NLP | Analysis of patient safety events related to look-alike/sound-alike | Look-alike/sound-alike errors involving opioid analgesics were more frequently associated with oxycodone | Preliminary research |
| (USA).[115] | reported by frontline staff. Data from Patient safety event (PSE) data in an academic healthcare system. |  |  | medication errors that occurred involving opioid analgesics. | products, particularly immediate release/extended release formulations and most occurred in the ordering stage of the medication process. Severe adverse effects were rare, but potentially life threatening. |  |
| Rifat 2019 (Bangladesh).[116] | Tweets containing keywords, primarily generic names of 17 opioid drugs. | 166,723 | NLP, recurrent Neural Networks, convolution neural network | Identification of opioid abuse from social media to reduce harm from opioid overdoses. | Convolutional recurrent neural network performed the best with an F1 score of 0.71. Out of 98 ADRs found from tweets, 50 could be mapped to Lowest Level Terms and 48 to Preferred Terms. Most adverse drug reactions related to codeine, fentanyl and tylenol. | Preliminary research. |
| Sarker 2019A (USA).[117] | Tweets using prescription and illicit opioid keywords. | 5,979 | NLP Random Forest | Automatic characterisation of opioid-related chatter to improve the current state of opioid toxico surveillance. | The classifier obtained an accuracy of 68.7%. The automatic classification experiments produced promising results, despite the small amount of annotated data, suggesting that automated, real-time opioid toxico surveillance may be a possibility with more annotated data. | Preliminary research |
| Sarker 2020 (USA). [118] | Reddit messages from 5 users who had mentioned opioid keywords (prescription or illicit) in their posts. | 4,288 posts | NLP | Analysis of opioid content in longitudinal data posted in a social media forum. | Longitudinal timelines revealed a variety of information including: their use of illicit and non-medical; use of prescription drugs; all five reported opioid addiction; social impacts of their drug use e.g. loss of employment; clinical consequences of addiction e.g., suicidality; challenges of opioid addiction and reoccurrence of use | Preliminary research |
| Vonkorff 2014 (USA).[119] | Chronic non-cancer pain patients receiving COT. Data from electronic medical records from Group Health. | 22,143 | NLP | Prevalence of prescription opioid abuse/overuse among COT patients. | Among 3663 NLP positive records, 77 % were manually validated as indicating opioid addiction, abuse or overuse. The prevalence rates for prescription opioid addiction, abuse or overuse were common among COT patients, with the highest rates observed among patients aged between 18-44. (12.3%) | Preliminary research. |

AUROC: area under the receiver operating characteristic curve; COT: chronic opioid therapy; EHR: electronic health records; NLP: Natural language processing; OUD: opioid use disorder; POU: problem opioid use

## APPENDIX 3.

Detailed summary of the four areas of AI application in opioid research

### 1: Risk prediction

Most of the studies reviewed focused on developing AI technology to support identification of factors that could predict the increased risk of developing an adverse opioid related outcome. The aim of this research being either to identify risks early and prevent resultant issues or, as a method of classification for opioid-dependent and long-term users. In this category, most of the AI tools researched focused on the development of prolonged opioid use following surgery. A range of factors were found to be predictive of prolonged use following surgery and included age; marital status; preoperative opioid use, medication, and haemoglobin; tobacco use; comorbidity of depression or diabetes and instrumentation. Other adverse outcomes explored were the risk of dependence, abuse, and overdose.

Some of the AI technology developed in this category was at a more advanced level of development with several researchers publishing their tools online as open access. However, to progress the validation and deployment of these at pace and scale within individual healthcare settings requires facilitation and guidance at a national level.

### 2: Surveillance and monitoring

Studies in this category investigated AI technology to improve surveillance and monitoring of misuse and illegal selling and to detect consequences that could result from opioid misuse. Most of the AI models in this category used natural language processing technology. The models ranged in their stage of development from preliminary research through to being available online as open access.

The purpose of the AI technology was to gather intelligence to support public health surveillance and prevent adverse consequences of opioid use. Adverse clinical consequences studied included HIV outbreaks triggered by opioid abuse and transition to injection drug use, suicidality, opioid overdose in opioid users from their social media posts, opioid-induced respiratory depression, and opioid induced constipation. In some of these studies the clinical consequence of opioid use had resulted in avoidable utilisation of healthcare resources, for example misdiagnosis of the cause of abdominal pain resulting in unnecessary surgery.[63]

Some studies in this category researched AI technology to classify different subgroups, estimate prevalence and identify the scale and location of illegal online selling. Illegal use of opioids, both selling and individual use, is a difficult area to tackle.

### 3: Pain management

Studies in this category explored pain management in various patient cohorts including adolescents from minority backgrounds and patients with depression concomitantly prescribed an antidepressant. Research also focused on patient characteristics that could determine opioid requirements post-surgery. AI models in this category ranged in their stage of development with some being at the preliminary stages of research, others required external validation, and some were available online as open access.

### 4: Patient support technology

This was the group with the least number of studies. Both studies in this category used smart phone technology. One study used a random forest algorithm to predict opioid craving or stress in the user through their movement as assessed by GPS.[88] The other study tested an AI enabled peer support platform that patients with OUD could use to support their recovery.[89] In both cases, further development of the model was being planned.

